# Rapid, high throughput, automated detection of SARS-CoV-2 neutralizing antibodies against native-like vaccine and delta variant spike trimers

**DOI:** 10.1101/2022.02.01.22270279

**Authors:** Narayanaiah Cheedarla, Hans P. Verkerke, Sindhu Potlapalli, Kaleb Benjamin McLendon, Anamika Patel, Filipp Frank, Gregory L. Damhorst, Huixia Wu, William Henry O’Sick, Daniel Graciaa, Fuad Hudaib, David N Alter, Jeannette Bryksin, Eric A. Ortlund, Jeanette Guarner, Sara Auld, Sarita Shah, Wilbur Lam, Dawn Mattoon, Joseph M Johnson, David H Wilson, Madhav V. Dhodapkar, Sean R. Stowell, Andrew S. Neish, John D. Roback

**Affiliations:** Department of Pathology and Laboratory Medicine, Emory University School of Medicine, Atlanta, GA 30322, USA; Department of Pathology, Brigham and Women’s Hospital, Boston, MA, USA; Department of Biochemistry, Emory University School of Medicine, Atlanta, GA 30322, USA; Department of Medicine, Division of infectious diseases, Emory University, Atlanta, GA 30322, USA; Rollins School of Public Health, Emory University, Atlanta, GA 30322, USA; Wallace H. Coulter Department of Biomedical Engineering, Georgia Institute of Technology and Emory University, Atlanta, GA, USA; Quanterix Corporation, 900 Middlesex Turnpike, Billerica, MA 01821; Department of Hematology/Medical Oncology, Emory University, Atlanta, GA

## Abstract

Traditional cellular and live-virus methods for detection of SARS-CoV-2 neutralizing antibodies (nAbs) are labor- and time-intensive, and thus not suited for routine use in the clinical lab to predict vaccine efficacy and natural immune protection. Here, we report the development and validation of a rapid, high throughput method for measuring SARS-CoV-2 nAbs against native-like trimeric spike proteins. This assay uses a blockade of hACE-2 binding (BoAb) approach in an automated digital immunoassay on the Quanterix HD-X platform. BoAb assays using vaccine and delta variant viral strains showed strong correlation with cell-based pseudovirus and live-virus neutralization activity. Importantly, we were able to detect similar patterns of delta variant resistance to neutralization in samples with paired vaccine and delta variant BoAb measurements. Finally, we screened clinical samples from patients with or without evidence of SARS-CoV-2 exposure by a single-dilution screening version of our assays, finding significant nAb activity only in exposed individuals. In principle, these assays offer a rapid, robust, and scalable alternative to time-, skill-, and cost-intensive standard methods for measuring SARS-CoV-2 nAb levels.

## Introduction

Levels of neutralizing antibodies (nAbs) against SARS-CoV-2 and other viruses predict vaccine efficacy and immune protection after natural infection^1-5^. In addition, the degree of protection from sterilizing immunity to prevention of severe disease correlates strongly with nAb levels at any given time post vaccination or infection^6^. Thus, the ability to reliably detect and quantify SARS-CoV-2 nAbs at scale is critical in the ongoing public health effort to reach population level protection in the face of waning immunity and a need for boosters^7^. In addition, the emergence of viral variants that escape neutralization by vaccine-induced antibodies underscores the importance of building efficient and reliable pipelines for nAb assay development as new variants are sequenced and rise to the level of interest or concern (VOI or VOC).

SARS-CoV-2 spike (S) protein is a large homotrimeric glycoprotein, which adopts a metastable prefusion conformation before its high affinity interaction with host-membrane associated angiotensin converting enzyme 2 (ACE-2)^8-10^. Native S protein forms two proteolytically cleaved extracellular subunits (S1 and S2), with S1 containing a specific 222 amino acid (AA) receptor binding domain (RBD) that binds to ACE-2^11-13^. Thus, S1 promotes receptor recognition and high affinity binding. The S2 subunit, in turn, drives membrane fusion through a fusion peptide (FP), two heptad repeat regions (HR1/2), and a transmembrane domain linked to the cytoplasmic tail^14^. To date, studies of neutralizing antibodies elicited by vaccination and natural infection as well as monoclonal antibody therapies have largely focused on antibodies that bind and inhibit interactions through SARS-CoV-2 RBD^15^. However, studies have also identified targets of neutralizing activity in SARS-CoV-2 S protein outside of the RBD, including regions in S2 proximal to the FP and HR2^16^. These findings were recently bolstered in a study by Garrett et al. using phage deep mutation scanning (Phage-DMS) to comprehensively interrogate immunodominant epitopes of antibodies in SARS-CoV-2 convalescent plasma as well as routes of antibody escape by the virus. This study independently identified non-RBD epitopes for neutralizing antibodies in FP and HR2^16^. Together these findings highlight the importance of closely approximating the native structure and domain organization of spike in any robust assay for SARS-CoV-2 neutralizing antibodies.

Current gold-standard assays for measuring nAbs against SARS-CoV-2 require live, replication-competent wild virus isolates or infectious molecular clones^17-18^. While these assays are important tools for research, they require a biosafety level 3 (BSL3) environment, are difficult to standardize, and are poorly suited for any scaled clinical application due to facilities, personnel, and safety requirements. A second tier of widely accepted nAb assays employ replication incompetent reporter viruses—commonly using backbones derived from either HIV or VSV—pseudotyped with SARS-CoV-2 Spike (S)^19-21^. These pseudovirus neutralization assays (PNAs) require only BSL2 working conditions and can be scaled for higher throughput. However, both live and pseudoviral assays require use and maintenance of living target cells, which introduces technical variability as well as regulatory complications to clinical testing operations that may seek to employ them. Furthermore, they are manual, labor-intensive assays with turn-around-times of several days. Finally, for lentivirus based PNAs, serum and plasma from patients receiving antiretroviral therapy or pre-exposure prophylaxis for HIV may contain inhibitors of pseudovirus activity non-specific to SARS-CoV-2^22^.

To address these limitations, we developed and validated a rapid, high throughput, automated blockade of ACE2 binding (BoAb) assay to quantify SARS-CoV-2 nAb activity against both vaccine (Wuhan-1) and delta (B.1.167.2) variant native-like trimeric spike proteins. This assay is performed on the ultrasensitive Quanterix-HDX platform, and is amenable to routine clinical use. We validated our BoAb by comparison to gold standard live virus and pseudovirus neutralization assays as well as clinically in samples from a cohort of SARS-CoV-2 exposed and vaccinated individuals collected during a serosurvey in the spring of 2021. In principle, our approach offers a rapid, adaptable, and scalable solution for detection of nAbs against any SARS-CoV-2 variant, against other viral pathogens, or against emerging viruses of pathogenic potential.

## Results

### Detection of SARS-CoV-2 neutralizing antibodies by novel automated assay for blockade of ACE-2 binding (BoAb)

The majority of SARS-CoV-2 neutralizing antibodies prevent viral entry by inhibiting the biochemical interaction between S protein and ACE-2. We therefore designed our assay to detect inhibition of this interaction by nAbs in patient samples or select inhibitors using SARS-CoV-2 spike conjugated beads as targets for binding by a biotinylated ACE-2 detector (**Fig. 1A)**. These reagents were then used in a custom three-step assay on the Quanterix HD-X platform (**Fig. 1B)**. To compare and quantify levels of neutralization from our BoAb assays, we engineered two primary readouts of the assay: an eight-point titration to identify the 50% inhibitory dilution or concentration (ID50 or IC50), and a single dilution readout calculated as a percentage of the maximum ACE-2 binding signal for a given spike target bead set. The latter approach was conceived as a potential screening tool for potent neutralizing antibodies while the former titering approach is appropriate for more rigorous comparisons among subjects or between candidate inhibitors (**Fig. 1C**).

**Figure 1.**
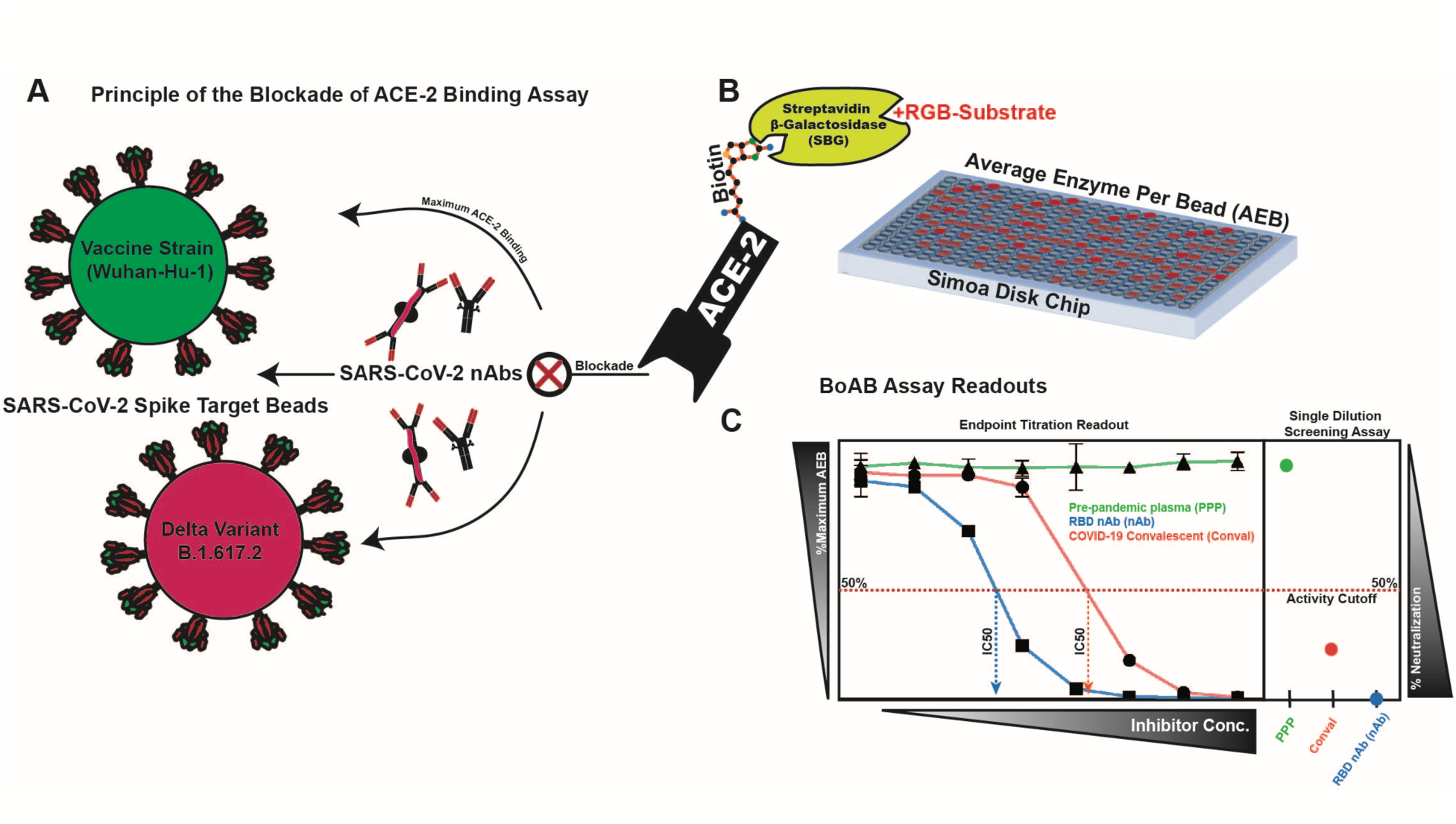
Blockade of ACE-2 Binding (BoAb) assay design. A schematic of our blockade of binding assay for SARS-CoV-2 neutralizing antibodies and its primary readouts: 50% inhibitory concentration (IC50) by titration or single dilution screening at a sample dilution of 1:50. (A) Detection of inhibitors of the ACE-2/SARS-CoV-2 spike interaction is achieved using an in-house purified and biotinylated human ACE-2 detector reagent. The ACE-2 binding signal is amplified by streptavidin-beta-galactosidase and a fluorescent RGB-Substrate. (B) The entire assay is automated and performed using single molecule array (SIMOA) technology on the Quanterix HD-X platform with a readout of average enzymes per bead (AEB). (C) Processed data from two assay readouts: titering for an IC50 (curves to the left) and screening for inhibition at a single sample dilution (right box).

### In-house generated vaccine strain and delta variant spike proteins adopt native trimeric structures and bind with high affinity to ACE-2

In order to present authentic, native-like spike targets for neutralizing antibody detection, we utilized a soluble, stabilized prefusion spike ectodomain construct originally designed in work by Hsieh et al^23^, but our plasmid DNA sequences are different for WT and Delta strain which were human codon optimized and synthesized at GenScript (Supplementary Text). This construct contains 6 stabilizing proline mutations in S2, which prevent the spontaneous and irreversible formation of a post-fusion state (**Fig. 2A)**. Spike targets for vaccine strain (Wuhan-Hu-1 GenBank: MN908947) and delta variant (B.1.617.2) were produced in human 293F cells and purified by affinity and size exclusion chromatography (SEC). A soluble, human IgG Fc chimera of ACE-2 was produced in a similar system before affinity and SEC. Purity of all in-house generated protein reagents was determined to be >95% using reducing SDS-PAGE **(Fig. S1)**. We studied the structures of purified proteins using negative stain electron microscopy NS-EM **(Fig 2 B, C)** with 2-dimensional class averaging and found that the proteins are formed in trimers. Further, we confirmed that our spike proteins adopted the expected homotrimeric structures with 3-fold symmetry at the apex and an expected tapering in the S1 to S2 transition (**Fig. 2D, E)**. Finally, we confirmed that our ACE-2 detector bound stably and with high affinity to both of these prefusion constructs using biolayer interferometry (**Fig. 2F, G)**. Both the vaccine strain and delta variant spike reagents bound the ACE2 detector at a similar steady state level and showed stable, slow dissociation rates. Together these data confirm the authentic structure of our spike reagents and their binding activity toward the ACE2 detector used in our BoAb assays.

**Figure 2.**
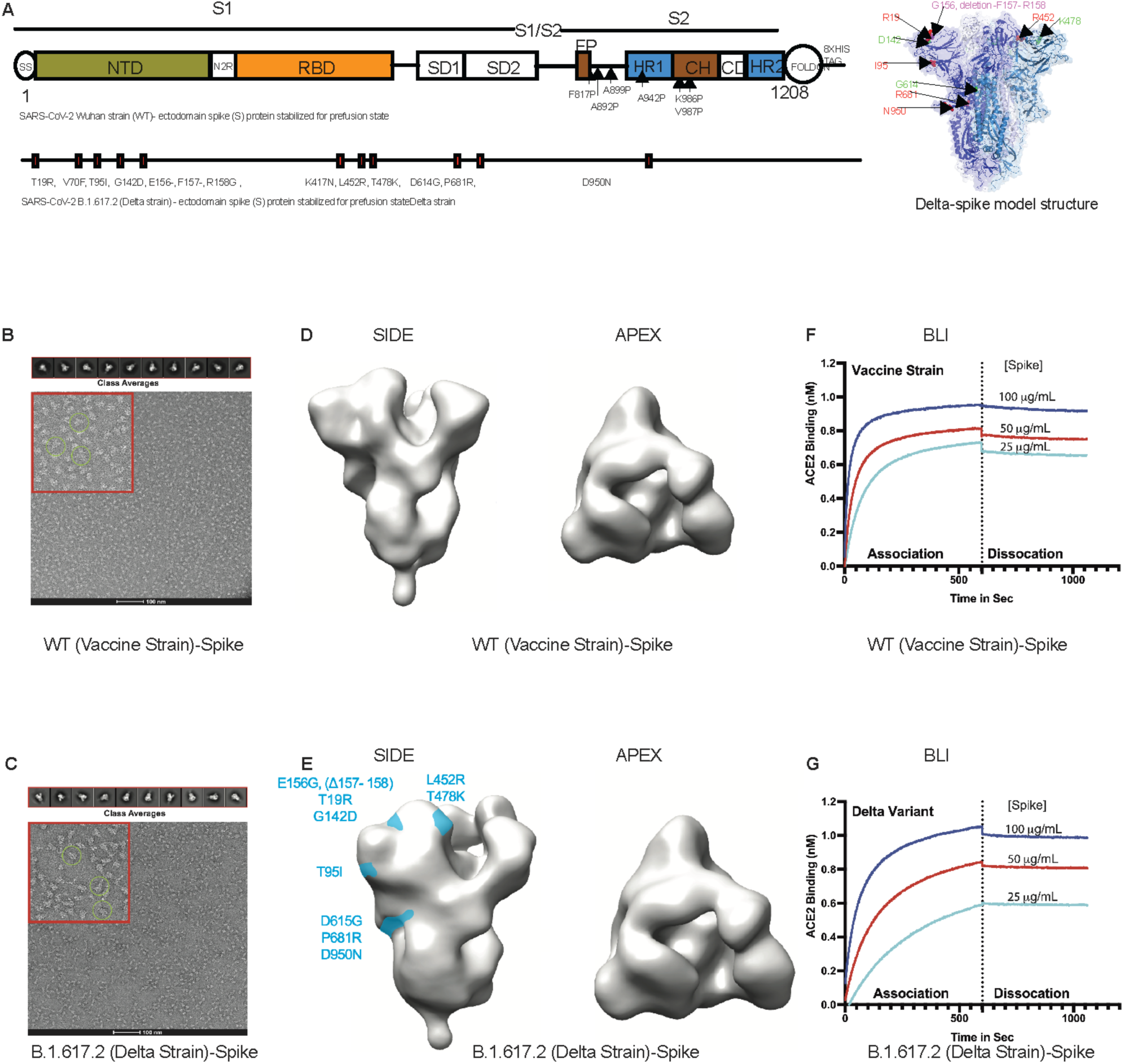
Development and validation of SARS-CoV-2 vaccine strain and delta variant prefusion ectodomain spike targets for use in blockade of ACE-2 binding assay (BoAb). (A) SARS-CoV-2 spike (S) ectodomain proteins of WT (Wuhan strain) and B.1.167.2 (Delta strain) for structural characterization and assay development, and a model construct of Delta S protein. **(B and C)** Raw negative stain electron micrographs for purified vaccine strain (B) and delta variant (C) trimers. Examples of native-like structures are encircled in the zoomed view, highlighted in a red cutout for each micrograph. 2D class averages of various trimer orientations derived from the raw micrographs are shown in the upper panel and used for the 3D reconstruction shown in Panels C and D. **(D)** 3D reconstruction from negative stain electron microscopy class averages of our purified vaccine strain trimeric ectodomain. **(E)** 3D reconstruction from negative stain electron microscopy class averages of our purified delta variant (B.1.617.2) trimeric ectodomain with significant amino acid substitutions mapped to the side view in light blue. **(F**,**G)** Biolayer interferometry analysis of each spike variant binding to an immobilized recombinant ACE-2 IgG Fc-chimera, the biotinylated form of which serves as the detector in our BoAb assay.

### Vaccine strain (Wuh-1) BoAb neutralizing activity correlates strongly with corresponding live virus and pseudovirus neutralization results

To determine the performance of our new test for SARS-CoV-2 neutralizing antibodies, we evaluated the correlation between titering results with the vaccine strain BoAb assay versus live virus and pseudovirus neutralization assays using plasma samples from patients vaccinated against COVID-19. Results from our vaccine strain assay showed strong correlation with results from a gold-standard live virus focus reduction neutralization test (FRNT) (**Fig. 3A)** as well as strong performance in a ROC analysis using the lowest reported log ID50 (1.17) as a cutoff for activity (AUC 0.94; P<0.0001) (**Fig. 3B and 3C)**. Similarly, our assay correlated strongly with vaccine strain pseudovirus neutralization activity, particularly in samples above the log ID50 limit of quantification (2.0) for our pseudovirus assay (R^2^ = 0.72; P<0.001) (**Fig. 3D)**. Using this pseudovirus LOQ as a cutoff for positivity, we performed a second ROC analysis and comparing vaccine strain BoAb activity in samples below and above the PNA LOQ. Our assay showed robust performance (ROC AUC 0.94; P<0.0001) with PNA results as a reference (**Fig. 3E and 3F)**. Our new assay also demonstrated strong correlation with levels of receptor binding domain (RBD) IgG, with samples containing higher levels of neutralizing antibodies as measured by BoAb showing significantly higher levels of RBD binding IgG (**Fig. 3G and 3H)**.

**Figure 3.**
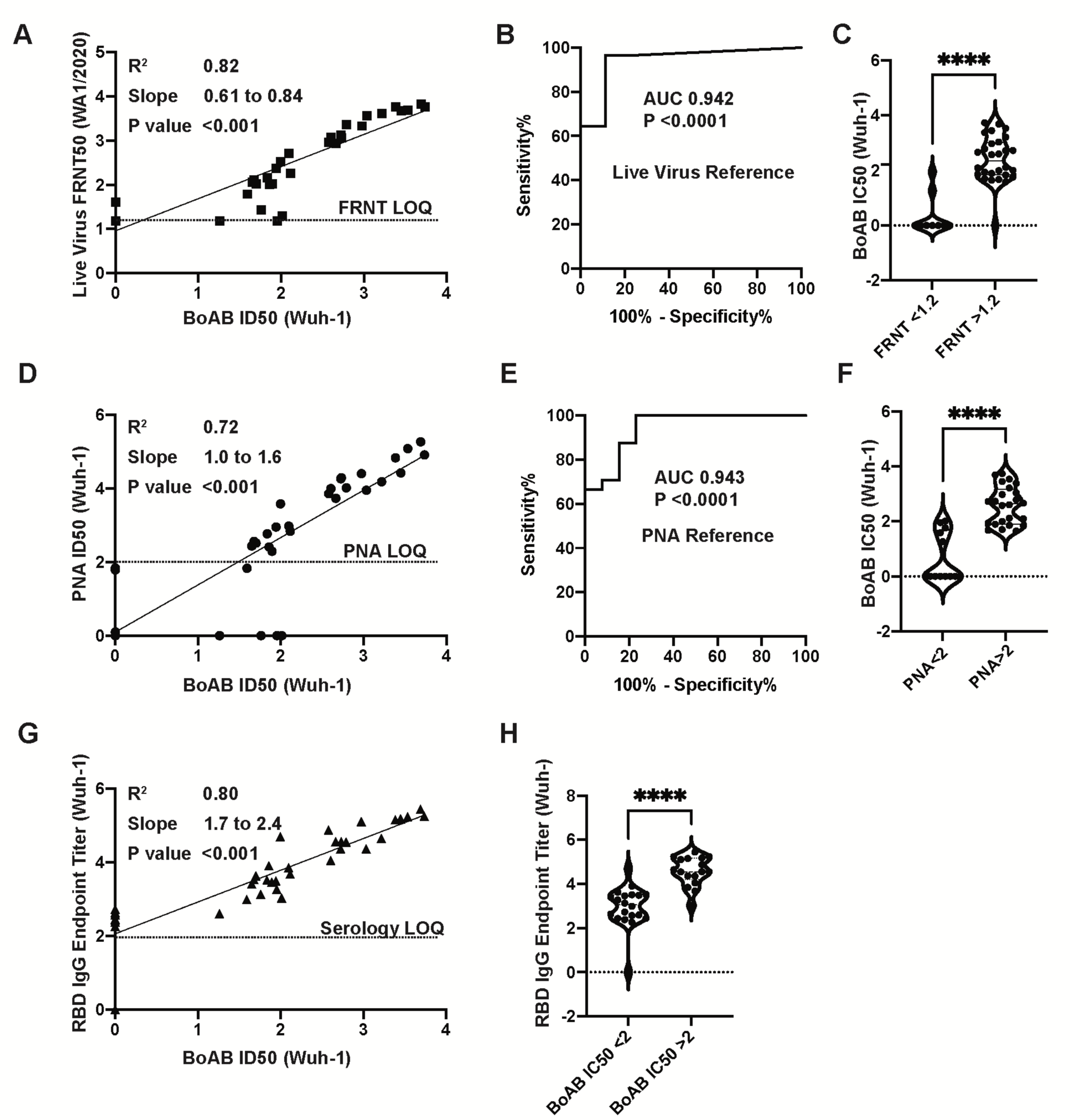
Correlation of vaccine strain BoAb IC50s with live virus and pseudovirus neutralization assays (LVN and PNA). (A) Linear regression analysis of vaccine lineage (Wuh-1 & WA1/2020) live virus 50% focus reduction neutralization activity (FRNT50) against ID50s in the vaccine strain BoAb. The absolute value of the log dilution factor at which a sigmoidal curve (fit to duplicate eight-point dilution series for each sample) crossed 50% is plotted for the BoAb assay. (B,C) Receiver operator characteristic (ROC) curve and categorical comparison of the vaccine strain BoAb using a live virus FRNT cutoff of 1.17 (representing a linear dilution of 1 in 15) as the reference standard for neutralizing activity. (D) Linear regression analysis of vaccine strain (Wuh-1 & WA1/2020) pseudovirus 50% inhibitory dilution (ID50) against ID50s in the vaccine strain BoAb. (E,F) Receiver operator characteristic (ROC) curve and categorical comparison of the vaccine strain BoAb using a pseudovirus neutralization ID50 of 2 (representing a linear dilution of 1 in 100) as the reference standard for neutralizing activity. (G) Linear regression analysis of vaccine strain (Wuh-1 & WA1/2020) receptor binding domain (RBD) specific IgG titers against IC50s in the vaccine strain BoAb. Values are plotted as in (A) using an optical density cutoff of 0.2 to quantify levels of binding antibodies. (H) Comparison of RBD IgG endpoint titers in samples with vaccine strain BoAb activity less than or greater than a log IC50 of 2. Statistical significance was evaluated by unpaired non-parametric t tests. ns=not significant, *P<0.05; **P<0.01; ***P<0.001; ****P<0.0001.

### Levels of delta variant (B.1.167.2) BoAb neutralization correlate strongly with corresponding live and pseudovirus virus neutralization results and accurately reflect patterns of escape from neutralizing antibodies

We next evaluated the performance of our assay for delta variant (B.1.167.2) neutralizing activity. We found a strong correlation and robust performance by ROC analysis for our new delta variant BoAb assay (R squared of 0.80; P<0.001) as compared to live delta variant neutralizing activity determined by gold standard FRNT assay (**Fig. 4A-C)**. ID50 results from our delta variant assay also correlated strongly with activity in our vaccine strain PNA and the corresponding live virus neutralizing antibody assay though, as expected, with a lower degree of correlation than that seen within strain (R squared of 0.66) (**Fig. 4D**). ROC analysis revealed a similar performance of the delta variant BoAb assay to the vaccine strain PNA assay using a vaccine strain PNA log ID50 cutoff of 2 for positivity **(Fig. 4E and 4F)**. Delta variant BoAb activity also correlated with vaccine strain RBD binding titers though to a lesser extent (**Fig. 4G)**. Finally, we evaluated the decrement between vaccine strain neutralizing antibody activity and delta variant activity, observed consistently in vaccinated individuals and postulated to be, at least in part, responsible for an increased frequency of delta variant breakthrough infections among vaccinated individuals^23^. Importantly, for each sample we found a similar pattern of decrement in vaccine strain and delta variant BoAb activity compared to live virus vaccine strain and delta variant FRNT results (**Fig. 4H and 4I)**. Together these data suggest that our delta variant BoAb assay correlates strongly with gold standard assays for neutralizing activity and may similarly detect deficits in delta variant specific activity observed among vaccinated individuals and those who experienced infection prior to the emergence of SARS-CoV-2 spike variants with the ability to escape nAbs.

**Figure 4.**
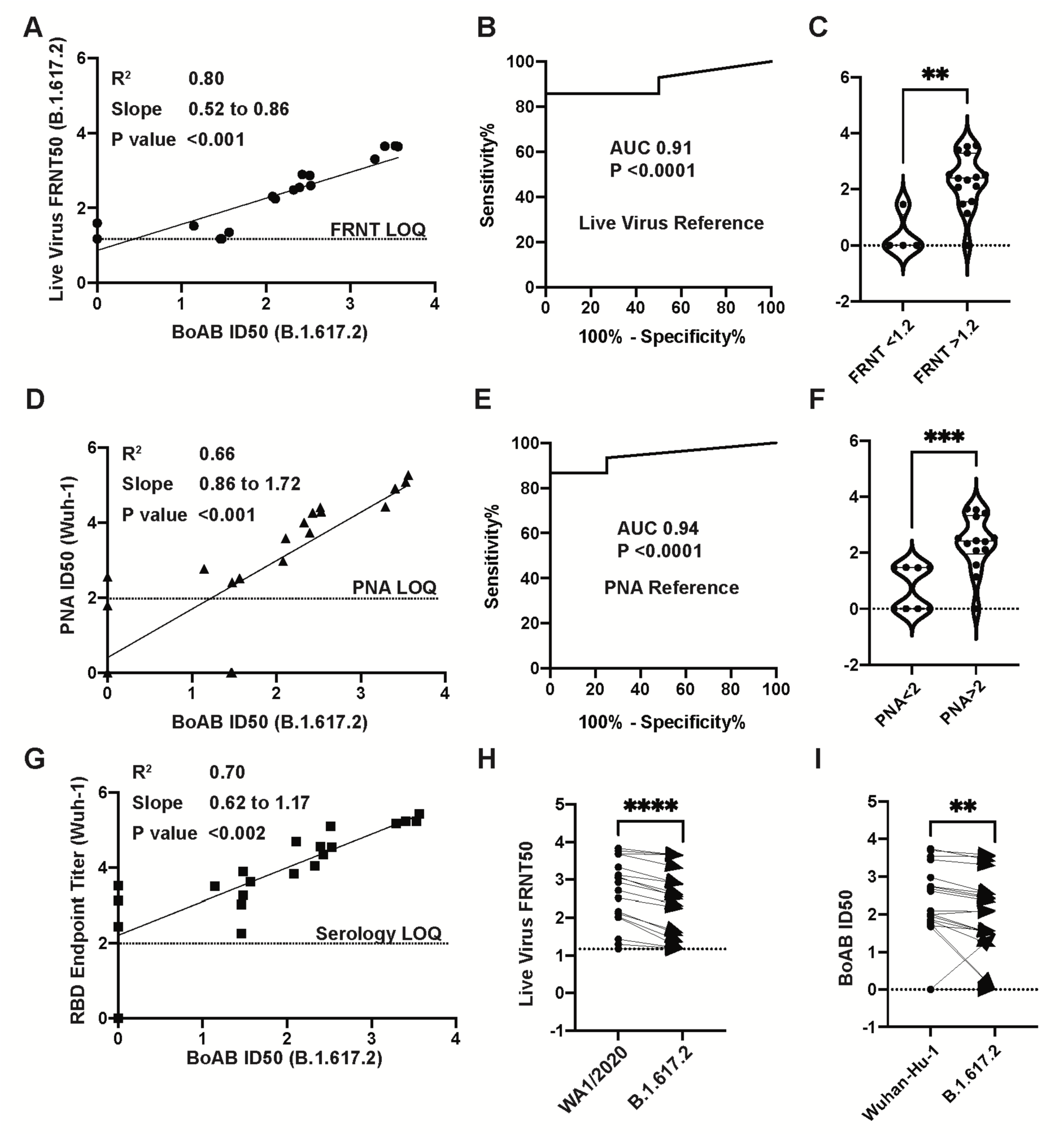
Correlation of delta variant BoAb ID50s with live virus and pseudovirus neutralization assays (LVN and PNA). A) Linear regression analysis of delta variant (B.1.617.2) live-virus FRNT 50% inhibitory concentration (ID50) against ID50s in the delta variant BoAb. (B,C) Receiver operator characteristic (ROC) curve and categorical comparison of the delta variant BoAb using a live virus FRNT cutoff of 1.17 (representing a linear dilution of 1 in 15) as the reference standard for neutralizing activity. (D) Linear regression analysis of vaccine strain (Wuh-1 & WA1/2020) pseudovirus 50% inhibitory concentration (ID50) against ID50s in delta variant (B.1.617.2) BoAb. (E,F) Receiver operator characteristic (ROC) curve and categorical comparison of the delta variant BoAb using a pseudovirus neutralization ID50 of 2 (representing a linear dilution of 1 in 100) as the reference standard for neutralizing activity. (G) Linear regression analysis of vaccine strain (Wuh-1) receptor binding domain (RBD) specific IgG titers against ID50 values in the delta variant BoAb. Values are plotted as in (A). (H,I) Paired comparison of live virus FRNT50 and BoAb ID50 values between vaccine strain lineage (Wuhan-Hu-1 or WA1/2020) and delta variant assays. Statistical significance was evaluated by paired non-parametric t tests ns=not significant, *P<0.05, **P<0.01, ***P<0.001,****P<0.0001.

### Screening for neutralizing antibody activity by single dilution BoAb among SARS-CoV-2 exposed patients

An ideal clinical screening test for SARS-CoV-2 neutralizing activity, in addition to being automated and well correlated with accepted standard assays, should not require limiting-dilution analysis which carries significant costs associated with skilled labor and resources. We therefore generated a single dilution screening test at a sample dilution that was well correlated with live-virus neutralizing activity (1:50) (**Fig. 5A)**. Next, we evaluated the correlation between single-dilution blockade of binding at 1:50 with quantitative spike IgG serology, also performed on the Quanterix platform using an EUA serology assay in samples from vaccinated individuals at various times after vaccination. We found a strong linear correlation between blockade of binding and levels of spike IgG in samples with spike specific IgG levels between 5 and 100 µg/mL. At higher concentrations, blockade of binding was saturated at 100% inhibition. Significant blockade was not detected in samples with less than 5 µg/mL of spike specific IgG (**Fig. 5B)**. Finally, the percentage neutralization at a 1:50 dilution was evaluated in a subset of samples from a serosurvey cohort collected in the Emory Healthcare system between January and March of 2021 among inpatients and outpatients who received a blood draw during the relevant encounter. Using available SARS-CoV-2 PCR testing data and serological results, we categorized patients into individuals more likely to have neutralizing activity at the time of sampling (exposed responders) and those unlikely to have nAb activity (unexposed, non-responders). Among 278 patients tested, we identified 115 who were serologically positive with evidence of SARS-CoV-2 exposure. Eighty-five patients were serologically negative without evidence of SARS-CoV-2 exposure at the time of the blood draw. All individuals who screened positive for significant neutralizing activity (>50% inhibition at a 1:50 dilution) in vaccine strain and delta variant single dilutions assays fell into the exposed responder category or had an unknown exposure status at the time of blood draw. Significantly more neutralizing activity was detected against the delta variant in this cohort, perhaps due to the fact that the circulating strain at the time (B1.167-Alpha)^24, 25^ carries many of the same spike mutations as the delta variant (**Fig. 5C)**. Together these data provide proof of concept for use and further validation of our multi-variant BoAb tests as screening tools in patients with evidence of SARS-CoV-2 exposure or vaccination.

**Figure 5.**
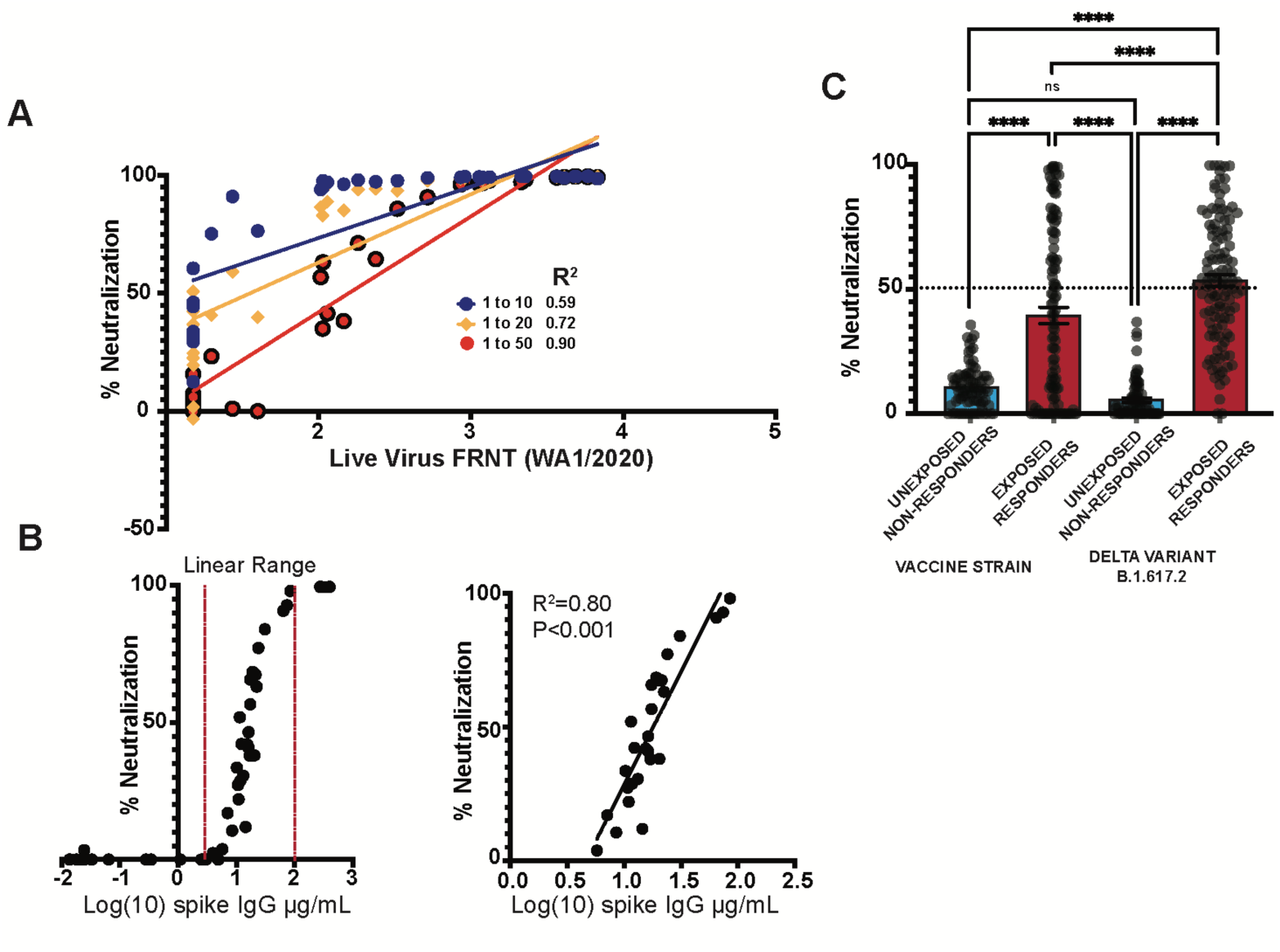
Detection of neutralizing antibodies by single dilution BoAb among SARS-CoV-2 exposed patients (A) Screening by linear regression analysis of various single plasma dilution activities (1 to 10, 1 to 20, and 1 to 50) for correlation with WA1/2020 live virus FRNT ID50. R squared values are shown for each regression. (B) Correlation of quantitative anti-spike IgG levels with 1:50 single dilution BoAb in all tested vaccinated individuals. Additionally, the correlation was plotted with a separate linear regression that was limited to samples with inhibition in the linear range (C). Comparison of single dilution delta and vaccine strain neutralizing antibody activity in patients with or without evidence of SARS-CoV-2 exposure and seroconversion from a serosurvey conducted in the spring of 2021 at Emory University Hospital Midtown. Statistical significance was evaluated by unpaired non-parametric t tests ns=not significant, *P<0.05, **P<0.01, ***P<0.001,****P<0.0001.

## Discussion

We report the development and validation of blockade of binding assays for the detection of nAbs against multiple SARS-CoV-2 variants. Results from our assays correlate strongly with established methods for nAb detection including live virus FRNT. Unlike these standard methods, our approach is completely automated and rapid, and does not require cell culture, BSL3 facilities, or extensive manual liquid handling. In addition, we employ spike antigens with native trimeric structure in our assays to capture the breadth of epitopes bound by vaccine and infection induced antibody responses. This latter point is particularly important with the roll out of boosters, which purportedly broaden the antibody response. An enhancement in neutralizing activity mediated by breadth of epitope specificity would be difficult to detect using subdomain and non-native spike targets.

Most viral infections are controlled by functional antibodies, like nAb, that block interactions between viral antigens and host receptors^1, 24-27^. In the case of SARS-CoV-2, spike protein interacts with the ACE-2 host cell receptor to enter host cells. Before interacting with host receptor, the spike antigen exists in a prefusion state. Conformational changes occur in the spike protein once it binds to ACE-2 receptor. Based on this finding, most assays were developed to detect nAbs in blood samples by incubating plasma with pseudo-typed viruses or live viruses prior to co-culture with target cells^28, 29^. It is well understood that viruses maintain the prefusion state of viral antigens (spike for SARS-CoV-2 and Env for HIV-1 and HIV-2) before interacting with host receptor, and most nAbs target the prefusion state of viral antigen^30-32^. Based on this model, we established an assay for the identification of nAbs against the prefusion state of SARS-CoV-2 spike protein that block interactions with human ACE-2 receptor. We have observed that the purified version of spike protein from Wuh-1 (WT) and Delta used in these studies maintained structures with one RBD up (opened confirmation) and two RBD down (closed confirmation) which was previously observed by Z Ke et al., when they studied spike structures on intact virion by cryo-EM images^9^. Thus, the purified spike proteins used in this assay maintain the prerequisite conformational state for the RBD-ACE2 interaction. Further, we observed that the spike proteins showed strong hACE-2 interaction by BLI.

For the initial analysis of BoAb assay, we used samples from Nooka et al.,^33^ who studied the neutralization determinants in myeloma patients. With this cohort, we observed that results in the BoAb assay strongly correlated with both live virus neutralization (Wuhan R^2^ = 0.82 and p=<0.001; Delta R^2^ = 0.80 and p=<0.001) and pseudovirus neutralization results (Wuhan R^2^ = 0.72 and p=<0.001; Delta R^2^ = 0.66 and p=<0.001). Interestingly, we also identified significant correlation of BoAb results with RBD endpoint titers for both Wuhan (p=<0.001) and Delta strains (p=0.002). These results agree with other studies in which RBD titers are strongly correlated with neutralization titers^34-36^. In addition, we determined that the BoAb assay showed sensitivity >94% against the Wuhan strain and >90% against Delta strain, both with 100% specificity. In general, neutralization against Delta strain was lower than for Wuhan strain, similarly to results from the BoAb assay.

We evaluated the utility of our BoAb assay in a second cohort of samples from a serological survey of patients requiring a blood draw in the Emory Healthcare system from January to March of 2021. We used available COVID-19 PCR testing results and combined serological status to categorize patients who were likely to have been exposed to SARS-CoV-2 and to have developed a humoral immune response against the virus. We hypothesized that this group of exposed responders had the highest pre-test probability in the cohort of harboring SARS-CoV-2 neutralizing activity and screened them using a 1:50 single dilution version of our BoAb assay. In agreement with our hypothesis, we observed strong neutralization against SARS-CoV-2 spike that was consistent with the patients’ exposure status. Therefore, our data is strongly aligned with correlations of antibody responses vs neutralization responses against SARS-CoV-2^37-39^. The rapid, automated, high-throughput BoAb assay described here may be useful for large-scale quantification of nAbs against SARS-CoV-2 variants for both clinical and investigational uses.

It should be noted that our study was limited by availability of gold-standard live-virus neutralizing antibody data and a need to directly correlate activity measured in our assay with known correlates of nAb activity in gold standard cell-based assays. While our data suggest that biochemical neutralization as measured by BoAb correlates well with results from these more established tests, additional work is needed to evaluate the implications of this association for vaccine efficacy and protection after natural infection.

## Methods

### Samples

Samples were sourced from various studies with IRB 00001663 at Emory University after obtaining the approval and consent from Institutional Review Board and samples (n=300) are tested on The Quanterix HD-X which uses single molecule arrays (SIMOAS) of femtoliter-sized reaction chambers etched into a disk to detect single enzyme labeled proteins. Our assay uses a 3-step ELISA that begins with an initial incubation of the sample and spike-conjugated magnetic bead reagent. Then the beads are separated magnetically and washed. Following a biotinylated ACE2 detector is added and incubated. A second magnetic separation and wash are performed. Followed by a streptavidin beta-galactosidase incubation and third wash. A resorufin ß-D-galactopyranoside substrate is added and the reaction chamber is sealed with oil. Any spike antigen that was able to attract a detector antibody will lead to the formation of an SBG/RGP fluorescent product that is counted digitally. The instrument can handle up to 288 replicates at a time.

### Pseudovirus neutralization assay (PNA)

Neutralization activities against SARS-CoV-2 WT (Wuh-1) and Delta (B.1.617.2) strains were measured in a single-round-of-infection assay with pseudo viruses, as previously described^33^. Briefly, to produce SARS-CoV-2 WT and Delta pseudoviruses, an expression plasmid bearing codon optimized SARS-CoV-2 full-length S plasmid (parental sequence Wuhan-1, Genbank #: MN908947.3) was co-transfected into HEK293T cells (ATCC#CRL-11268) with plasmids encoding non-surface proteins for lentivirus production and a lentiviral backbone plasmid expressing a Luciferase-IRES-ZsGreen reporter, HIV-1 Tat and Rev packing plasmids (BEI Resources) and pseudoviruses harvested after 48 hours of post transfection and performed titration. Pseudoviruses were mixed with serial dilutions of plasma or antibodies and then added to monolayers of ACE-2-overexpressing 293T cells (BEI Resources), in duplicate. 24 hours after infection, cells were lysed, luciferase was activated with the Luciferase Assay System (Promega), and relative light units (RLU) were measured on a synergy Biotek reader.

### Protein expression and purification

Trimeric SARS-CoV-2 Spike (Wuh-1 and Delta B.1.617.2) as well as Angiotensin Converting Enzyme-2 (ACE-2)-IgFC chimera proteins were produced by transfection in FreeStyle 293-F cells using plasmids (MN908947 for Wuhan-1 and Delta strains, and NM_021804.3 for hACE-2). Briefly, FreeStyle 293F cells were seeded at a density of 2E6 cells/ml in Expi293 expression media and incubated with shaking on at 37°C and 127 rpm with 8% CO2 overnight. The following day, 2.5E6 cells/ml were transfected using ExpiFectamine™ 293 transfection reagent (ThermoFisher, cat. no. A14524) according to the manufacturer protocol. Transfected cells were then incubated with orbital shaking for 4-5 days at 37 °C,127 rpm, 8% CO2. Supernatants containing secreted trimeric ectodomains were collected by centrifugation at 4,000xg for 20 minutes at 4°C. Clarified supernatants were then filtered using a 0.22 µm stericup filter (ThermoFisher, cat.no. 290-4520) and loaded onto pre-equilibrated affinity columns for protein purification. The SARS-CoV-2 Spike trimer and ACE-2 proteins were purified using His-Pur Ni-NTA resin (ThermoFisher, cat.no. 88221) and Protein-G Agarose (ThermoFisher, cat.no. 20399) respectively. Briefly, His-Pur Ni-NTA resin was washed twice with PBS by centrifugation at 2000xg for 10 min. The resin was resuspended with the spike-trimer supernatant and incubated for 2 hours on a shaker at RT. Gravity flow columns were then loaded with supernatant-resin mixture and washed (25mM Imidazole, 6.7 mM NaH_2_PO_4_.H_2_O and 300 mM NaCl in PBS) four times, after which the protein was eluted in elution buffer (235 mM Imidazole, 6.7 mM NaH_2_PO_4_.H_2_O and 300 mM NaCl in PBS). Eluted protein was dialyzed against PBS using Slide-A-lyzer Dialysis Cassette (ThermoScientific, Cat# 66030) and concentrated using 100 kDa Amicon Centrifugal Filter Unit, at 2000 g at 4°C. The concentrated protein eluate was then run and fractionated on a Sepharose 600 (GE Healthcare) column on an Akta™Pure (GE Healthcare). Fractions corresponding to the molecular weight of each protein were pooled and concentrated as described above. Proteins were quantified by BCA Protein Assay Kit (Pierce) and quality was confirmed by SDS-PAGE (supplementary Figure 1).

### ACE-2 protein expression and purification

The soluble ACE-2 IgFC chimera was expressed as described above. Clarified supernatants were diluted 1:1 with binding buffer before loading on a protein g gravity flow column, pre-equilibrated with 10 ml of binding buffer (Pierce cat.no.21011). Columns were washed with 20 ml of binding buffer, and the protein was eluted in 40 ml of elution buffer. Following elution, samples were first neutralized to pH 7.5 using 1 M Tris, pH 9.0. Eluted protein was dialyzed against 50mM Tris (pH7.5), 150mM NaCl using a Slide-A-lyzer Dialysis Cassette (ThermoScientific, Cat# 66030) and concentrated using 50 kDa Amicon Centrifugal Filter Unit, at 2000 g at 4°C. Size exclusion chromatography and quality control was performed on the concentrated protein as described above.

### Assessment of Spike-ACE-2 binding by biolayer interferometry

6x His-tagged spike was diluted to 50 µg/mL in PBS before immobilization on nickel NTA biosensors (fortebio). Association of ACE-2 was monitored using and OctetRED96e instrument (fortebio) in 2-fold dilutions series starting at 100 µg/mL for 600s followed by dissociation in PBS for 500s. Tips were regenerated using 10mM glycine and regenerated in 10 mM NiCl2 before re-loading with equivalent concentrations of spike.

### Generation of detector and conjugated beads

ACE-2 detector biotinylation and spike bead conjugation were performed per the Quanterix Homebrew Detection Antibody Biotinylation and Bead Conjugation Protocols.

1. *ACE-2 Biotinylation* Briefly, ACE-2 was buffer exchanged using Amicon filtration into Quanterix biotinylation reaction buffer prior to mixing at 1mg/mL with a 40x challenge ratio of NHS-PEG4-biotin for 30 minutes at room temperature. Cleanup of the biotinylated detection reagent was achieved by a further round of amicon filtration following recovery in biotinylation reaction buffer and determination of protein concentration. A final detector concentration of 0.5µg/mL was used in the assay.
2. *Spike conjugation with magnetic beads* Paramagenetic beads were activated after washing with bead conjugation buffer using 9ug EDC (10mg/mL) in a final bead volume of 300 µl containing 4.2E8 beads for 30 minutes at 4 °C with rocking. Following activation, beads were washed with Bead Conjugation Buffer and 300 µl cold spike at 0.2mg/mL to the beads followed by incubation at 4 °C with rocking for 2 hours. Beads were then washed and blocked for 45 minutes at room temperature, followed by a final wash and resuspension in 300 µl bead diluent. Spike capture beads were stored at 4 °C until use in the assay.

### Negative Stain sample preparation, data collection and data analysis

Spike protein was diluted to 0.05mg/ml in PBS prior to grid preparation. A 3 µL drop of diluted protein applied to previously glow-discharged, carbon coated grids for ∼60 sec, blotted and washed twice with water, stained with 0.75% uranyl formate, blotted and air dried. Between 25-35 images were collected on Talos L120C microscope (Thermo Fisher, Waltham, MA) at 73,000 magnification and 1.97 Å pixel size. Relion-3.1 was used for particle picking, 2D classification and 3D reconstruction [PMID: 23000701].

### Statistical Analysis

GraphPad PRISM version 9 was used to perform the statistical analysis. Correlation between the assays was performed by Pearson r correlation method and linear regression analysis. All statistical tests were two-sided, unless otherwise noted, and statistical significance was assessed at the *P<0.05, **P<0.01, ***P<0.001, ****P<0.0001 and further details are provided in the figure legend where analysis was performed.

## Data Availability

All data produced in the present work are contained in the manuscript

## Acknowledgments

We would like to thank Cato, Lee from Eric Ortlund lab for help in processing SEC for protein purification and we also acknowledge Amit Joshi, Todd Glynn and Danielle Svancara from Quanterix team for their tremendous support while developing assay on HDX instrument. We acknowledge BEI Resources for providing plasmid (NR-52309) and 293T-ACE-2 cell line (NR-52511). The Robert P. Apkarian Integrated Electron Microscopy Core (IEMC) at Emory University is subsidized by the School of Medicine and Emory College of Arts and Sciences. Additional support is provided by the Georgia Clinical & Translational Science Alliance of the National Institutes of Health under award number UL1TR000454. We would like to acknowledge Connie Arthur and Cheryl Meier for their fabulous support with serosurvey specimen processing.

## Authors Contributions

N.C., H.V., A.S.N., and J.D.R. contributed to the acquisition, analysis, and interpretation of the data, S.P., K.B.M., W.H.Os., and F.H. interpretation of data, A.P., and F.F. involved in the interpretation of the NS-EM images, G.D., H.W., J.B., J.G., S.A., S.S., and M.V.D provided samples from Emory Healthcare, D.N.A., E.O., W.L., S.R.S., A.S.N and J.D.R. contributed to the data analysis, and A.S.N., and J.D.R received grants from NIH/NCI, 1 U54 CA260563-01: Immune regulation of COVID-19 infection in cancer and autoimmunity and Emory CURE grant from Emory University, Atlanta, USA.

---

**Supplementary Figure 1.**
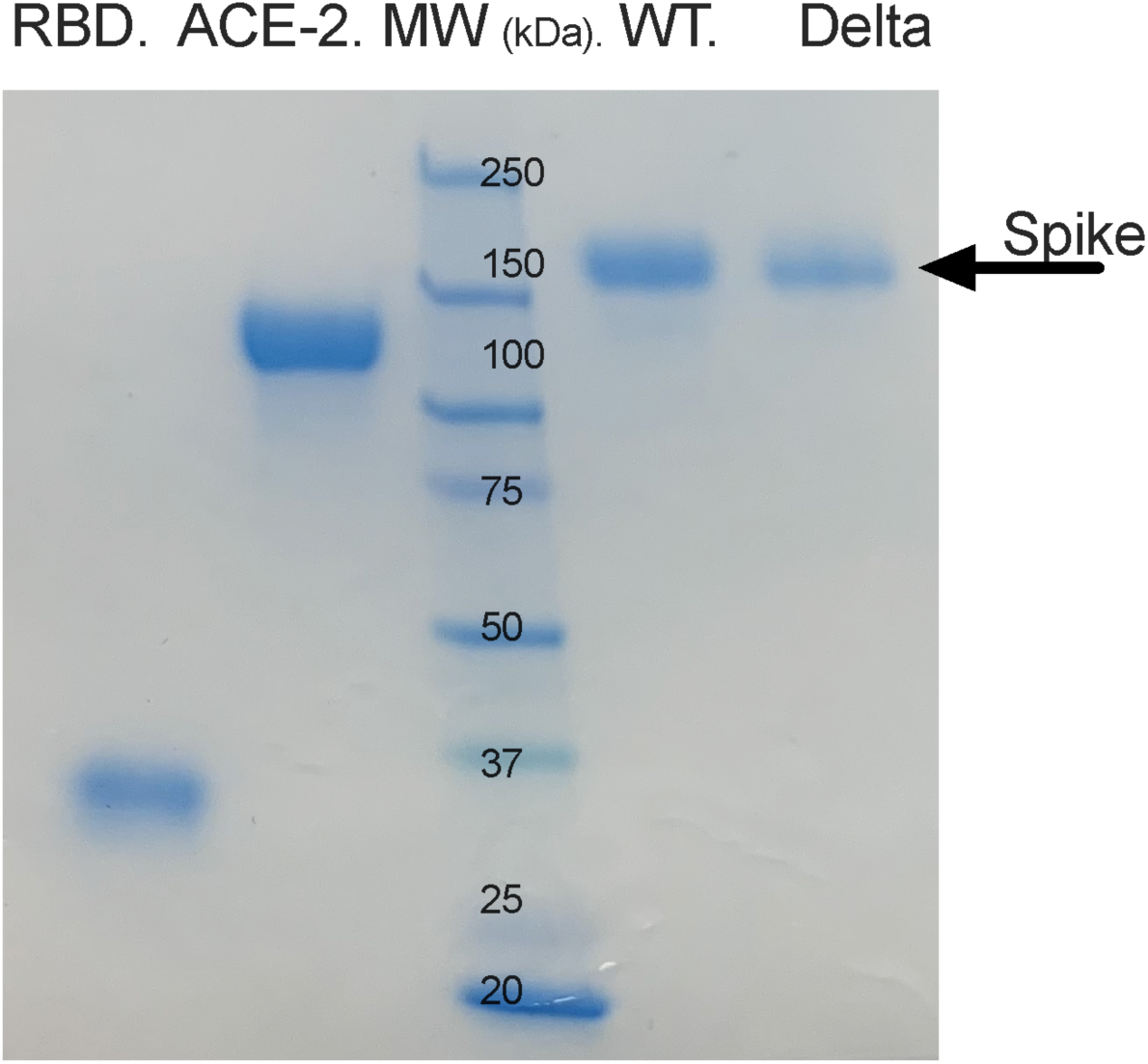
SDS-PAGE for proteins from SARS-CoV-2 RBD, WT, Delta and hACE-2 expressed in Expi293 cells and purified by affinity and size exclusion chromatography.

